# Differences in COVID-19 Vaccination and Experiences among Patients with Hypertension in Colombia and Jamaica during the COVID-19 Pandemic

**DOI:** 10.1101/2024.03.25.24304855

**Authors:** Jacqueline P Duncan, Siyi Geng, Carene Lindsay, Trevor Ferguson, Katherine Mills, Jose Patricio Lopez-Lopez, Hua He, Paola Lanza, Makeda Williams, Veronica Tutse-Tonwe, Mabel Reyes, Alfonso Campo, Allison Marshall, Patricio Lopez-Jaramillo, Marshall K Tulloch-Reid

## Abstract

**Background:** COVID-19 vaccination and shielding targeted hypertensive patients in low and middle income countries. We describe the COVID-19 experiences of hypertensive patients in Colombia and Jamaica and discuss factors associated with vaccine acceptance.

**Methods:** A cross-sectional study was conducted between December 2021 and February 2022 in 4 randomly selected primary care clinics in Colombia and 10 primary care clinics in Jamaica. Participants in Colombia were randomly selected from an electronic medical record. In Jamaica consecutive participants were selected on clinic days for non-communicable diseases. Interviewer-administered questionnaires were conducted by telephone.

**Results:** 576 participants were recruited (50% Jamaica; 68.5% female). Jamaica’s participants were younger (36% vs 23% <60 years) and had a lower proportion of persons with “more than high school” education (17.2% vs 30.3%, p=0.011). Colombia’s participants more commonly tested positive for COVID-19 (24.2% vs 6.3%, p<0.001), had a family member or close friend test positive for COVID-19 (54.5% vs, 21.6%; p<0.001), experienced loss of a family member or friend due to COVID-19 (21.5% vs 7.8%, p<0.001) and had vaccination against COVID-19 (90.6% vs 46.7%, p<0.001). Fear of COVID-19 (AOR 2.71, 95% CI 1.20-6.13) and residence in Colombia (AOR 5.88 (95% CI 2.38-14.56) were associated with COVID-19 vaccination. Disruption in health services affecting prescription of medication or access to doctors was low (<10%) for both countries.

**Conclusion:** Health services disruption was low but COVID-19 experiences such as fear of COVID-19 and vaccine acceptance differed significantly between Colombia and Jamaica. Addressing reasons for these differences are important for future pandemic responses.

## Introduction

Coronavirus disease 2019 (COVID-19) was declared a public health emergency of international concern on 30 January 2020. Non-pharmaceutical interventions such as border closures, curfews, mandatory face masks, and hand sanitization were implemented in countries to stave off waves of the pandemic (1). The impact of non-pharmaceutical interventions (NPI) disrupted multiple economic sectors, social norms and individual’s livelihoods, disproportionately affecting the fragile economies of low and middle income countries (LMICs) (2). This was further exacerbated by delays in access to vaccines – resulting in a prolonged use of the NPIs for disease management in these settings which slowed economic recovery in these settings (3).

In Jamaica and Colombia, the health sector experienced direct and indirect consequences of the COVID-19 response which likely contributed to the excess deaths observed during the pandemic (4). Direct effects of the pandemic included disruption of medication supply chains and health services as non-COVID care was cancelled or delayed (5,6). Potential indirect impacts include increase in body mass index due to changes in diet and physical activity, exposure to domestic violence and an increase in mental health disorders such as depression and anxiety (7). These would have special impacts in persons living with chronic diseases such as hypertension (8–10).

Hypertension was identified as one of the risk factors for COVID-19 infection, severe COVID-19 and death (11,12). As a result, these patients were prioritized for vaccination and shielding as part of the COVID-19 response worldwide. Differences in vaccine acceptance between and within countries were recorded including variations by ethnic groups (13,14). Jamaica for instance had approximately 26.3% of the population fully vaccinated by December 31, 2022 compared to 71.1% of the population in Colombia (15). Low vaccine acceptance has also been recorded for ethnic minorities in the United States of America and the United Kingdom (14).

The COVID-19 experiences of persons residing in LMICs during the pandemic are not well documented despite the burden of non-communicable diseases (NCDs) and the propensity for sustained disruptions in health care delivery and supply chain management. As part of a needs assessment for the Caribbean and South America Team-based Strategy to Control Hypertension (CATCH) Study conducted during the pandemic, we were able to explore the COVID-19 experiences of hypertensive patients in Colombia and Jamaica and examined factors associated with vaccine acceptance. Conducting research in these two settings with significant differences in vaccination uptake also provided a unique opportunity to understand reasons for vaccine hesitancy that may be relevant to developed countries such as the USA where vaccine uptake in Black and Hispanic communities was low.

## Methods

### Study Setting

Colombia and Jamaica are upper middle income countries in the Latin America and Caribbean (LAC) region. Persons with hypertension are managed in primary care centers that are strategically placed based on population size and density. Both countries offer a mix of public and private health services. Colombia’s health care system is characterized by access to a network of public and/or private providers that provide care based on enrollment in a selected Benefit Plan Administrative Entity (EAPB). Outpatient management of hypertensive patients occur at Service Provider Institutions and information is captured on an electronic medical record which is not always interconnected between EAPB.

Jamaica’s health services are also a public – private mix with half of the population accessing services in the public sector. In the public sector, more than 300 community-based health centers offer primary care services to persons with hypertension at no cost. While stand-alone databases capture limited administrative information, the health information system is largely paper-based.

### The CATCH Study Design

The CATCH Study is a cluster randomized trial to test the implementation and effectiveness outcomes of implementing and scaling up a team-based care strategy for BP control in Colombia and Jamaica. As part of the needs assessment for CATCH, a cross-sectional study was conducted in 4 randomly selected primary care clinics in Colombia and 10 primary care clinics in Jamaica between December 2021 and February 2022.

In Jamaica, consecutive patients were selected from clinic attendees on clinic days for non-communicable diseases. Colombia’s patients were randomly selected from an electronic medical record system. Stratified sampling was used to ensure selection of similar numbers of urban and rural participants, as well as representation of both sexes and all age groups. Interviewer-administered questionnaires collected information on hypertension knowledge, lifestyle changes, medication adherence and COVID-19 experiences (testing positive for COVID-19, COVID-19 vaccination, disruption in health services impacting prescription refills). All interviews were conducted by telephone. Informed consent was obtained by telephone or during the face-to-face encounter (depending on the method of recruitment) before the start of the interview. Copies of the consent form were provided to participants via Whatsapp, email or regular mail based on their preferences. For those who were recruited on-site physical copies of the consent form were also provided.

The study protocol was approved by the Tulane University Institutional Review Board (USA), Institutional Bioethics Committee of Universidad de Santander (Colombia) and the University of the West Indies (UWI) Mona Campus Research Ethics Committee (Jamaica).

### Measurements

A questionnaire was developed to inform the intervention for hypertension control in the CATCH Study. Validated questions were sourced from approved toolboxes and, since the survey was conducted during the COVID-19 pandemic, data was captured on COVID-19 experiences including vaccination. The questionnaire was piloted among 20 patients with hypertension (10 Colombia and 10 Jamaica) at two primary care clinics in Colombia and one primary care clinic in Jamaica. Surveys were administered in English and Spanish in Jamaica and Colombia respectively.

The outcome variable “COVID-19 vaccination” was measured by asking the question: “Have you been vaccinated against COVID-19?” with dichotomized responses “yes” or “no”. Independent variables included location (urban/rural based on clinic location), COVID-19 infection, complacency and country of residence.

COVID-19 infection was determined by answering “yes” to either of the following questions “*Since the COVID-19 pandemic began, have you experienced any of the following life events: a) Testing positive for COVID-19? b) Family member or close friend testing positive for COVID-19?”*

To assess “complacency” we asked respondents if “fear of contracting coronavirus if you leave the house” prevented them from seeing a doctor or getting their prescriptions.

### Sample size

We determined that a minimum of 72 participants per primary subgroup (male/female and urban/rural status) will be needed to detect a 20% response proportion with a 95% confidence interval from 10%-30%. Based on sample size calculations, assuming stratification by sex and urban/rural status, we propose inclusion of 288 participants per country.

### Statistical Analysis

Analyses were done using SAS 9.4 (SAS Institute, Cary, NC). Descriptive statistics such as mean and standard deviation for continuous variables, and frequency and percentage for categorical variables are provided for the study sample. Data were weighted to account for differences in the age, sex and urban/rural area distribution of the study participants. Weights were determined by assessing the joint distribution of age groups, sex, and residential areas within the respective target population of each country from clinic registers. In analyses that included participants from both countries, combined weights were applied.

Bivariate analyses were used to examine differences in COVID-19 experiences (testing positive, family member/friend positive, COVID-19 vaccination and disruption of services impacting prescription service) and urban/rural status. Logistic regression was used to examine factors associated with COVID-19 vaccination among hypertensive patients in both settings as we explored reasons for differences in uptake. A *P* < .05 was considered statistically significant.

## Results

Two hundred and eighty-eight people with hypertension were surveyed in each country (total of 576 participants). The majority of participants were female (68.5%), ≥ 60 years old (70.9%), employed/retired (69.7 %) and primarily from urban areas (68.1%). Colombian participants were older (mean age 66.5 vs 62.5 years old) and had higher proportion of persons with “more than high school education” (30.3% vs 17.2%, p=0.011). Higher levels of being unemployed (14.0% vs 6.0%, p<0.001) and “never married” (34.0% vs 5.5%, p<0.001) was reported in Jamaica.

### Health Service Disruption

Table 2 shows the proportion of participants that experienced service disruption in Jamaica and Colombia based on geographical location (urban/rural). Few participants (<10%) were unable to see a doctor or get prescriptions due to clinic or pharmacy closure, curfews, having less time with a doctor, loss of health insurance or unaffordability of healthcare (Table 2). However, urban-rural differences in COVID-19 experiences were observed as the negative impact of these NPIs on prescription or medical doctor access was more common for participants in rural areas. For example, in rural areas 13.0% and 10.4% participants in Colombia and Jamaica respectively reported being “unable to see doctor/get prescription due to facility closure” compared to 1.6% and 2.1% in urban areas.

**Table 1:** Baseline characteristics of participants (N=576) by country of residence (Jamaica and Colombia)

| **Characteristic** | **All Participants**<br>**N=576, n (%)<sup>a</sup>** | **Jamaica**<br>**N=288, n (%)<sup>a</sup>** | **Colombia**<br>**N=288, n (%)<sup>a</sup>** | **P-value** |
| --- | --- | --- | --- | --- |
| **Age - years (mean (SD))** | 64.6 (11.4) | 62.5 (10.2) | 66.5 (12.2) | 0.001 |
| **Age** |  |  |  | 0.006 |
| <60 | 234 (29.1) | 128 (36.0) | 106 (22.9) |  |
| 60-69 | 226 (30.3) | 106 (31.0) | 120 (29.6) |  |
| ≥ 70 | 116 (40.6) | 54 (32.9) | 62 (47.5) |  |
| **Sex** |  |  |  | 0.217 |
| Male | 274 (31.5) | 130 (34.4) | 144 (28.9) |  |
| Female | 302 (68.5) | 158 (65.6) | 144 (71.1) |  |
| **Marital Status** |  |  |  | <.0001 |
| Never Married | 105 (18.8) | 92 (34.0) | 13 (5.5) |  |
| Married/ Common Law | 323 (50.3) | 130 (40.6) | 193 (58.9) |  |
| Widowed | 65 (17.0) | 27 (12.4) | 38 (21.1) |  |
| Divorced | 25 (5.8) | 14 (4.9) | 11 (6.6) |  |
| Other | 58 (8.0) | 25 (8.1) | 33 (7.9) |  |
| **Location** |  |  |  | 0.367 |
| Urban | 290 (68.1) | 146 (66.0) | 144 (69.9) |  |
| Rural | 286 (31.9) | 142 (34.0) | 144 (30.1) |  |
| **Comorbidities** |  |  |  |  |
| Diabetes | 168 (26.4) | 116 (38.3) | 52 (15.9) | <.001 |
| High cholesterol | 288 (52.7) | 159 (58.9) | 129 (47.1) | 0.029 |
| Depression | 31 (5.3) | 9 (3.1) | 22 (7.3) | 0.064 |
| Anxiety | 29 (5.0) | 10 (3.7) | 19 (6.1) | 0.287 |
| Kidney Disease | 18 (3.6) | 4 (1.1) | 14 (5.8) | 0.001<sup>b</sup> |
| Stroke | 30 (4.9) | 25 (7.7) | 5 (2.5) | <.001<sup>b</sup> |
| Heart Attack/Heart Disease | 36 (6.1) | 8 (2.7) | 28 (9.1) | 0.008 |
| Overweight/Obesity | 269 (42.9) | 98 (31.9) | 171 (52.7) | <.001 |
| **Education Category** |  |  |  | 0.011 |
| Less than High school | 215 (37.9) | 121 (43.2) | 94 (33.2) |  |
| High school | 227 (37.9) | 121 (39.6) | 106 (36.5) |  |
| More than High school | 134 (24.1) | 46 (17.2) | 88 (30.3) |  |
| **Occupation Category** |  |  |  | <.001 |
| Employed/retired | 411 (69.7) | 214 (76.1) | 197 (64.0) |  |
| Unemployed | 66 (9.7) | 42 (14.0) | 24 (6.0) |  |
| Housewife | 65 (15.8) | 5 (1.5) | 60 (28.5) |  |
| Disabled | 34 (4.7) | 27 (8.4) | 7 (1.4) |  |
| **Multi-morbidity** |  |  |  | 0.053 |
| Yes (≥2 conditions) | 269 (44.1) | 142 (49.5) | 127 (39.2) |  |
| No | 307 (55.9) | 146 (50.5) | 161 (60.8) |  |

<sup>a</sup> The sample sizes and actual frequency numbers are unweighted but means (SDs) or percentages are weighted.
<sup>b</sup> Fisher's exact test method, weights are rounded up to the nearest non-zero integer for test.

**Table 2.** Impact of COVID-19 on healthcare access and utilization by urban/rural status in Jamaica and Colombia.

| Health Service Disruption | All participants<br>N=576 | Jamaica |  |  | Colombia |  |  |
| --- | --- | --- | --- | --- | --- | --- | --- |
|  |  | Urban<br>N=146 | Rural<br>N=142 | P-value<sup>b</sup> | Urban<br>N=144 | Rural<br>N=144 | P-value<sup>b</sup> |
| Unable to see doctor/get prescription due to facility closure, n (%)<sup>a</sup> | 35<br>(4.9) | 2<br>(1.6) | 12<br>(13.0) | <.001<sup>†</sup> | 4<br>(2.1) | 17<br>(10.4) | <.001<sup>c</sup> |
| Unable to get prescription due to pharmacy closure, n (%)<sup>a</sup> | 23<br>(3.5) | 0 | 7<br>(8.2) | <.001<sup>†</sup> | 5<br>(3.8) | 11<br>(7.0) | 0.069<sup>c</sup> |
| Less time to see a doctor or get medications due to curfew restrictions, n (%)<sup>a</sup> | 49<br>(7.5) | 8<br>(5.5) | 6<br>(3.9) | 0.548 | 6<br>(3.8) | 29<br>(19.5) | <.001 |
| Did not seek care due to fear of contracting coronavirus if you leave the house, n (%)<sup>a</sup> | 217<br>(39.6) | 9<br>(6.6) | 10<br>(7.5) | 0.802 | 87<br>(61.1) | 111<br>(79.8) | 0.004 |
| Loss of insurance or government assistance, n (%)<sup>a</sup> | 8<br>(1.2) | 3<br>(1.2) | 1<br>(0.8) | 0.647<sup>†</sup> | 2<br>(0.7) | 2<br>(1.0) | 0.653<sup>c</sup> |
| Trouble affording medications or healthcare costs, n (%)<sup>a</sup> | 28<br>(4.5) | 14 (7.7) | 6<br>(4.2) | 0.216 | 2<br>(1.6) | 6<br>(4.8) | 0.058<sup>c</sup> |

<sup>a</sup> The sample sizes and actual frequency numbers are unweighted, but percentages are weighted.
<sup>b</sup> p-value for rural/urban difference
<sup>c</sup> Fisher's exact test method, weights are rounded up to the nearest non-zero integer for test

### COVID-19 Experiences

Overall, 15.8% participants reported testing positive for COVID-19 (24.2% Colombia and 6.3% Jamaica, p<0.001). Colombia’s participants more commonly experienced having a family member or close friend test positive for COVID-19 (54.5% vs, 21.6%; p<0.001), loss of a family member or friend due to COVID-19 (21.5% vs 7.8%, p<0.001) and having vaccination against COVID-19 (90.7% vs 46.7%, p<0.001). However, 44.4% of Jamaica’s participants were willing to be vaccinated if available (Table 3). Testing positive for COVID-19 was more common for urban participants in Colombia compared to rural participants (28.1% vs 15.3%, p=0.025) but no other significant differences in these COVID-19 experiences were recorded based on location (urban/ rural) in each country (data not shown).

**Table 3:** COVID-19 infection and vaccination by country of residence (Jamaica and Colombia)

|  | **All**<br>**Participants**<br>**N=576** | **Jamaica**<br>**N=288** | **Colombia**<br>**N=288** | **P value** |
| --- | --- | --- | --- | --- |
| Tested positive for COVID-19, n (%)<sup>a</sup> | 96 (15.8) | 22 (6.3) | 74 (24.2) | <.001 |
| Family member or close friend tested positive for COVID-19, n (%)<sup>a</sup> | 225 (39.0) | 71 (21.6) | 154 (54.5) | <.001 |
| Loss of a family member or close friend due to COVID-19, n (%)<sup>a</sup> | 85 (15.1) | 23 (7.8) | 62 (21.5) | <.001 |
| Vaccinated against COVID-19, n (%) | 397 (70.1) | 133 (46.7) | 264 (90.7) | <.001 |
| If not vaccinated, willingness to be vaccinated if available, n (%)<sup>a</sup> | 86 (42.4) | 74 (44.4) | 12 (32.2) | 0.363 |
| Vaccinated against COVID-19 or willingness to be vaccinated if available, n (%)<sup>a</sup> | 483 (82.9) | 207 (70.6) | 276 (93.7) | <.001 |

<sup>a</sup> The sample sizes and actual frequency numbers are unweighted, but percentages are weighted.

Fear of contracting coronavirus was more common among Colombian participants (68.8% vs 6.5%, p<0.001) and also more common among persons who were vaccinated or willing to be vaccinated against COVID-19 if available (47.0% vs 4.7%, p<0.001)).

### Factors associated with COVID-19 vaccination status

The characteristics of the sample by vaccination status are presented in Table 4. Three hundred and ninety-seven participants were vaccinated (90.7% Colombia, 46.7% Jamaica, p<0.001) with vaccinated participants being older than unvaccinated participants (mean age 65.4 vs 62.8 years old), more frequently married or in a common law relationship (56.8% vs 35.2%) and with higher rates of overweight/obesity (47.4% vs 32.3%). Other demographic characteristics and comorbidities were similar in both groups.

**Table 4:** Baseline characteristics of participants (N=576) in Jamaica and Colombia by COVID-19 Vaccination status.

| Characteristic | All Participants<br>N=576 | Vaccination<br>N=397 | No Vaccination<br>N=178 | P-value |
| --- | --- | --- | --- | --- |
| Age – years (mean (SD)) | 64.6 (11.4) | 65.4 (11.6) | 62.8 (10.6) | 0.042 |
| Age, n (%)<sup>a</sup> |  |  |  | 0.343 |
| <60 | 234 (29.1) | 153 (27.1) | 81 (33.7) |  |
| 60-69 | 226 (30.3) | 160 (30.1) | 65 (30.4) |  |
| ≥ 70 | 116 (40.6) | 84 (42.8) | 32 (35.8) |  |
| Sex, n (%)<sup>a</sup> |  |  |  | 0.082 |
| Male | 274 (31.5) | 188 (29.0) | 86 (37.5) |  |
| Female | 302 (68.5) | 209 (71.0) | 92 (62.5) |  |
| Marital Status, n (%)<sup>a</sup> |  |  |  | <.001 |
| Never Married | 105 (18.8) | 45 (11.3) | 60 (36.6) |  |
| Married/ Common Law | 323 (50.3) | 250 (56.8) | 73 (35.2) |  |
| Widowed | 65 (17.0) | 48 (19.5) | 16 (10.9) |  |
| Divorced | 25 (5.8) | 15 (5.1) | 10 (7.5) |  |
| Other | 58 (8.0) | 39 (7.3) | 19 (9.8) |  |
| Location, n (%)<sup>a</sup> |  |  |  | 0.341 |
| Urban | 290 (68.1) | 192 (66.8) | 98 (71.2) |  |
| Rural | 286 (31.9) | 205 (33.2) | 80 (28.8) |  |
| Comorbidities, n (%)<sup>a</sup> |  |  |  |  |
| Diabetes | 168 (26.4) | 104 (23.7) | 64 (32.9) | 0.065 |
| High cholesterol | 288 (52.7) | 196 (52.2) | 91 (53.6) | 0.798 |
| Depression | 31 (5.3) | 20 (4.7) | 11 (6.7) | 0.423 |
| Anxiety | 29 (5.0) | 21 (5.0) | 8 (5.2) | 0.927 |
| Kidney Disease | 18 (3.6) | 16 (3.8) | 2 (3.1) | 0.310<sup>b</sup> |
| Stroke | 30 (4.9) | 12 (3.5) | 18 (8.2) | 0.072 |
| Heart Attack/Heart Disease | 36 (6.1) | 28 (6.1) | 8 (5.9) | 0.948 |
| Overweight/Obesity | 269 (42.9) | 203 (47.4) | 65 (32.3) | 0.006 |
| Education Category, n (%)<sup>a</sup> |  |  |  | 0.265 |
| Less than High school | 215 (37.9) | 144 (37.3) | 70 (39.2) |  |
| High school | 227 (37.9) | 150 (36.3) | 77 (41.9) |  |
| More than High school | 134 (24.1) | 103 (26.4) | 31 (18.9) |  |
| Occupation Category, n (%)<sup>a</sup> |  |  |  | <.001 |
| Employed/retired | 411 (69.7) | 285 (68.9) | 126 (71.9) |  |
| Unemployed | 66 (9.7) | 40 (7.4) | 26 (15.2) |  |
| Housewife | 65 (15.8) | 56 (20.7) | 8 (4.1) |  |
| Disabled | 34 (4.7) | 16 (3.0) | 18 (8.8) |  |
| Multi-morbidity, n (%)<sup>a</sup> |  |  |  | 0.040 |
| Yes (≥2 conditions) | 269 (44.1) | 178 (40.5) | 90 (52.3) |  |
| No | 307 (55.9) | 219 (59.5) | 88 (47.7) |  |

<sup>a</sup> The sample sizes and actual frequency numbers are unweighted, but percentages are weighted.
<sup>b</sup> Fisher's exact test method, weights are rounded up to the nearest non-zero integer for test.

The association between COVID-19 vaccination and socio-demographic features as well as COVID-19 experiences is shown in Table 5. In a multivariable model adjusted for socio-demographic factors, fear of COVID-19 (AOR 2.71, 95% CI 1.20-6.13) and country of residence (AOR 5.88 (95% CI 2.38-14.56) for Colombians compared to Jamaica) increased the odds of COVID-19 vaccination. Both were independently associated with vaccination against COVID-19 (Table 5).

**Table 5.** Logistic Regression Model of Factors associated with COVID-19 Vaccination of Hypertensive patients in Colombia and Jamaica.

| Characteristic | Crude OR<br>(95% CI) | Adjusted OR <sup>a</sup> (95% CI) |
| --- | --- | --- |
| Age (per 1 increase) | 1.02 (1.00-1.04) | 1.01 (0.99-1.03) |
| Sex |  |  |
| Male | Ref | Ref |
| Female | 1.47 (0.95-2.27) | 1.33 (0.75-2.35) |
| Education |  |  |
| Less than High school | Ref | Ref |
| High school | 0.91 (0.54-1.53) | 0.79 (0.39-1.61) |
| More than High school | 1.47 (0.79-2.72) | 0.97 (0.44-2.15) |
| Location |  |  |
| Urban | Ref | Ref |
| Rural area | 1.23 (0.80-1.88) | 1.36 (0.81-2.29) |
| COVID-19 infection |  |  |
| No COVID-19 infection or<br>infection of family member | Ref | Ref |
| COVID-19 infection or infection<br>of family member | 2.75 (1.73-4.37) | 1.41 (0.82-2.42) |
| Complacency |  |  |
| No Fear of COVID-19 infection | Ref | Ref |
| Fear of COVID-19 infection | 9.38 (5.22-16.84) | 2.71 (1.20-6.13) |
| Country |  |  |
| Jamaica | Ref | Ref |
| Colombia | 11.06 (5.85-20.90) | 5.88 (2.38-14.56) |

<sup>a</sup> Adjustment for age, sex, education, rural/urban area, COVID-19 infection, fear of COVID-19 infection and country.

## Discussion

Our study found significantly higher levels of COVID-19 infection, fear of COVID-19 and COVID-19 vaccine acceptance in patients with hypertension in Colombia compared to Jamaica. Since hypertension is a risk factor for COVID-19 infection and severe COVID-19, these patients were targeted for immunization against COVID-19 across LAC (16). Colombia and Jamaica were the first countries to receive COVID-19 vaccines in the Americas and the Caribbean respectively through the COVID-19 Vaccine Global Access (COVAX) initiative (17). A meta-analysis examining vaccine acceptance in LAC reported high levels of vaccine acceptance (pooled prevalence of 78%, 95% CI: 74.0%–82.0%) but only one Caribbean country, Puerto Rico, was included in the pooled analysis (18).

In our study, COVID-19 vaccination was significantly higher in Colombia compared to Jamaica (90.6% versus 46.7%, p<0.001). These findings are similar to the differences in COVID-19 vaccine acceptance in the general population of both countries. By the end of the study period (February 1 2022), 61.2% and 22.0% of the general population in Colombia and Jamaica respectively were fully vaccinated (19,20). To date, Jamaica has the second lowest level of COVID-19 vaccination in the Caribbean with approximately 30% of the population fully vaccinated.

Low acceptance of COVID-19 vaccine in LMICs has been attributed to the global inequity in vaccine distribution and vaccine hesitancy (21). Conceptual frameworks such as the “three Cs model of vaccine hesitancy” indicate that complacency or low perceived risk of vaccine preventable diseases as well as convenience (e.g. availability and acceptability) and confidence (or trust) are the main factors that influence vaccine hesitancy (22,23). Of note, 44.4% of Jamaica’s participants were willing to be vaccinated, if available, suggesting that “convenience” or accessibility of COVID-19 vaccines may have contributed to the low uptake despite the establishment of fixed vaccination sites across the island.

Meta-analyses show that female sex, younger age, unemployment and lower educational attainment are associated with vaccine hesitancy (13,24). Our analysis did not show any associations between vaccination and age, sex, education, or location (urban/rural). However, in multivariate models, vaccination was associated with fear of COVID-19 (adjusted OR 2.71 (1.20-6.13) and fear of COVID-19 was significantly higher in Colombia compared to Jamaica (68.8 vs 6.5%). Although COVID-19 infection (self or family member) is reported as a predictor for vaccine acceptance in some studies, we did not find an association between vaccination and history of COVID-19 infection when models adjusted for socio-demographic variables and country of residence (25,26).

Understanding and targeting the reasons for complacency or low perceived threat of COVID-19 infection in LMICs such as Jamaica and among ethnic minorities in HIC should be a priority for improving vaccine acceptance. Inadequate access to diagnostic testing resulting in under-diagnosis of COVID-19 infections and deaths is one possible explanation for “complacency” in Jamaica. Many LMIC had a shortage of rapid tests due to global inequities in rapid testing and excess deaths during the pandemic have been attributed to undiagnosed COVID-19 and inadequate diagnostic capacity(27).

COVID-19 antigen rapid testing was introduced in Colombia and Jamaica in July 2020 and in September 2020 respectively (28,29). Colombia boasts having one of the highest COVID-19 testing rates in LAC with 33.9 million tests (or 666,009 tests per million people) done up to April 2022 (30,31). COVID-19 testing was widely available and by March 2022, the number of COVID-19 tests ranged from 13,000 to 39,000 per day. In Jamaica, COVID-19 testing targeted hospitalized persons as well as symptomatic persons seeking care at public health facilities. Rapid testing was available in the private sector but the high costs made home testing unaffordable for most of the population. Approximately 320,000 tests per million people were done in Jamaica by March 2022 and 14,000 rapid tests were done between January 30 and February 5, 2022 (32). This difference in access to testing in Jamaica and Colombia may also explain the finding that Colombia’s participants more commonly tested positive (24.2% vs 6.3%, p<0.001), had a family member or close friend test positive (54.5% vs, 21.6%; p<0.001) and experienced losing a family member or friend to COVID-19 (21.5% vs 7.8%, p<0.001).

Low complacency or perceived threat of COVID-19 and experiences with COVID-19 infection (self and family member testing positive) may also be due to the severity of the pandemic in each country. By February 1, 2022, Colombia which has a population of 52.1 million people was experiencing its 3^rd^ wave of COVID-19 with a cumulative total of 5,901,715 cases and 134,551 deaths, a crude mortality rate of 258 deaths per 100,000 people (33,34). In contrast, Jamaica has a population of 2.9 million people and recorded 124,806 cases, 2663 deaths, crude mortality rate of 92 deaths per 100,000 population (35). Jamaica was also experiencing its 3^rd^ wave. The dominant strain was Omicron BA1 in both countries (30). Based on the higher crude mortality rate in Colombia, it is possible that Colombia experienced more severe waves of COVID-19 with confounding by age, co-morbidities or other factors.

Although not well documented, the pandemic’s impact on health care delivery is likely to be greater in LMICs as they bear the burden of NCDs (70% NCD deaths, 85% premature deaths due to NCDs) and lack resilient health systems (6,36). Our study found that most participants (>90%) in Colombia and Jamaica did not experience reduced access to prescription medications or to a doctor in primary care due to service disruptions or curfews. Medication adherence was generally not affected by any of the COVID-19 experiences measured in our study. However, urban/rural differences were observed. For example, accessing doctors or prescription medications due to clinic or pharmacy closure was at least 5 times more common for participants in rural areas compared to urban areas. High levels of service disruption due to NPIs are reported in other studies with important impacts on hypertension care. For example, a 2020 World Health Organization (WHO) survey of 163 member states showed that 53% countries surveyed in the pandemic had disruption of services for hypertension treatment (37). Other studies reported reduced access to prescription medications and medication adherence as well as delayed diagnoses for persons with NCDs(38,39). Shah et al(40) showed that uncontrolled hypertension increased from 15% to 19% during the pandemic in the USA. A more detailed assessment of the impact of NPIs on health care delivery is needed to avoid interruption in care and adverse health outcomes for patients with hypertension and other chronic diseases.

Our study has important strengths as it provides information on COVID-19 experiences from a relatively large sample of hypertensive patients in two LAC countries. Although LMICs are disproportionately affected by NCDs such as HTN and COVID-19, few studies record vaccine acceptance and other COVID-19 experiences in these countries. Understanding the experiences of persons with hypertension during the COVID-19 pandemic will provide important information for planning the response to future shocks to the health system including epidemics like COVID-19 and Chikungunya. Studies documenting patient experiences in different LMIC settings are of particular value in understanding how local factors – including health care systems – impacted these experiences. While there is documentation of differences in vaccine uptake in minority populations in the USA there are limited data on these experiences in many of the source populations to which these ethnic groups belong. This provides additional insight into factors that influenced this practice.

However, this study has some limitations. Some documented predictors of vaccination (e.g. trust in government and source of vaccine information) and important COVID-19 experiences (e.g. use of Telehealth, access to other diagnostic and screening tests) were not captured in this study. Although we report low levels of service disruption affecting prescribed medications, the quality and nature of health services as well as subsequent health outcomes for persons with hypertension need to be assessed. Vaccine acceptance and recall of COVID-19 experiences are time-dependent and may have changed given the dynamic nature of the pandemic response. Finally, the findings may not be generalizable to other countries in LAC, but provides important information for improving vaccine acceptance in future pandemics.

## Conclusion

Access to rapid testing and severity of the pandemic are possible explanations for the different experiences with COVID-19 infection, complacency and COVID-19 vaccine acceptance in Colombia and Jamaica. Low perceived threat of COVID-19 (or complacency) and inadequate access to COVID-19 vaccination (convenience) may have contributed to the significantly lower COVID-19 vaccination uptake in Jamaica compared to Colombia but did not explain the significant country differences in uptake in these setting. Ensuring equitable distribution and access to rapid testing may be important for improving vaccine uptake in future pandemics or public health emergencies. Greater emphasis should be placed on community engagement which is necessary for building trust and strengthening implementation of national responses. A comprehensive review of the COVID-19 response and experiences in LMIC can provide valuable information for public health policy and planning.

## Disclosure

The authors declared no potential conflicts of interest with respect to the research, authorship, and/or publication of this article. No copyrighted materials were used in this article.

## Data Availability

All data produced in the present study are available upon reasonable request to the authors

## Acknowledgments

This work was supported by the National Heart, Lung, and Blood Institute (4UH3HL152373). We would like to thank the project staff as well as the members of the Health Departments and participants in Colombia and Jamaica who facilitated the conduct of this study.

## Disclaimer

The views expressed in this manuscript are those of the authors and do not necessarily represent the views of the National Heart, Lung, and Blood Institute; the National Institutes of Health; or the U.S. Department of Health and Human Services.

## Data availability

The data underlying this article will be shared on reasonable request to the corresponding author.

## Notes

**Funding:** This work was supported by the National Heart, Lung, and Blood Institute (4UH3HL152373).

**Conflicts of interest:** None.

### Competing Interest Statement

The authors have declared no competing interest.

### Funding Statement

Funding: This work was supported by the National Heart, Lung, and Blood Institute (4UH3HL152373). Disclaimer: The views expressed in this manuscript are those of the authors and do not necessarily represent the views of the National Heart, Lung, and Blood Institute; the National Institutes of Health; or the U.S. Department of Health and Human Services.

